# Towards a multimodal neuroimaging-based risk score for mild cognitive impairment by combining clinical studies with a large (N>37000) population-based study

**DOI:** 10.1101/2024.03.12.24303873

**Authors:** Elaheh Zendehrouh, Mohammad S. E. Sendi, Anees Abrol, Ishaan Batta, Reihaneh Hassanzadeh, Vince D. Calhoun

**Affiliations:** Tri-institutional Center for Translational Research in Neuroimaging and Data Science: Georgia State University, Georgia Institute of Technology, Emory University Atlanta, GA; Department of Electrical and Computer Engineering at Georgia Institute of Technology, Atlanta, GA; Departments of Psychology and Computer Science, Georgia State University, Atlanta, GA; Harvard Medical School and McLean Hospital, Boston, MA

**Keywords:** Mild cognitive impairment, multimodal neuroimaging, brain risk score, gray matter, functional network connectivity

## Abstract

Alzheimer’s disease (AD) is the most common form of age-related dementia, leading to a decline in memory, reasoning, and social skills. While numerous studies have investigated the genetic risk factors associated with AD, less attention has been given to identifying a brain imaging-based measure of AD risk. This study introduces a novel approach to assess mild cognitive impairment MCI, as a stage before AD, risk using neuroimaging data, referred to as a brain-wide risk score (BRS), which incorporates multimodal brain imaging. To begin, we first categorized participants from the Open Access Series of Imaging Studies (OASIS)-3 cohort into two groups: controls (CN) and individuals with MCI. Next, we computed structure and functional imaging features from all the OASIS data as well as all the UK Biobank data. For resting functional magnetic resonance imaging (fMRI) data, we computed functional network connectivity (FNC) matrices using fully automated spatially constrained independent component analysis. For structural MRI data we computed gray matter (GM) segmentation maps. We then evaluated the similarity between each participant’s neuroimaging features from the UK Biobank and the difference in the average of those features between CN individuals and those with MCI, which we refer to as the brain-wide risk score (BRS). Both GM and FNC features were utilized in determining the BRS. We first evaluated the differences in the distribution of the BRS for CN vs MCI within the OASIS-3 (using OASIS-3 as the reference group). Next, we evaluated the BRS in the Alzheimer’s Disease Neuroimaging Initiative (ADNI) cohort (using OASIS-3 as the reference group), showing that the BRS can differentiate MCI from CN in an independent data set. Subsequently, using the sMRI BRS, we identified 10 distinct subgroups and similarly, we identified another set of 10 subgroups using the FNC BRS. For sMRI and FNC we observed results that mutually validate each other, with certain aspects being complementary. For the unimodal analysis, sMRI provides greater differentiation between MCI and CN individuals than the fMRI data, consistent with prior work. Additionally, by utilizing a multimodal BRS approach, which combines both GM and FNC assessments, we identified two groups of subjects using the multimodal BRS scores. One group exhibits high MCI risk with both negative GM and FNC BRS, while the other shows low MCI risk with both positive GM and FNC BRS. Moreover, in the UKBB we have 46 participants diagnosed with AD showed FNC and GM patterns similar to those in high-risk groups, defined in both unimodal and multimodal BRS. Finally, to ensure the reproducibility of our findings, we conducted a validation analysis using the ADNI as an additional reference dataset and repeated the above analysis. The results were consistently replicated across different reference groups, highlighting the potential of FNC and sMRI-based BRS in early Alzheimer’s detection.

## 1. Introduction

Alzheimer’s disease (AD) is a neurodegenerative condition in individuals over 65 that affects the brain and causes memory, cognitive, and behavioral impairment. AD patients may experience disorientation, language difficulties, loss of independence, and personality changes as the illness worsens (Knopman et al., 2021). AD does not presently have a cure, but there are treatments and lifestyle modifications that can help control symptoms and enhance quality of life for both patients and their caregivers (Yiannopoulou and Papageorgiou, 2013; Yu and Tan, 2015). Given the substantial impact AD has on patients, their families, and society as a whole, in addition to the fact that there is no effective treatment, a preventative approach is essential. Prevention methods might both delay the development and lower the prevalence of dementia (Crous-Bou et al., 2017; Sindi et al., 2015). Before the onset of AD, some individuals may experience mild cognitive impairment (MCI), a stage characterized by cognitive decline greater than expected for one’s age but not severe enough to interfere significantly with daily activities. As such, finding an MCI risk score that can identify the individual with a high risk of developing AD is vital for the prevention approach.

Much previous research has focused on the genetic risk of AD (Stocker et al., 2018). While some genetics research has shown that some genes can enhance a person’s risk of the illness, having a gene variation does not guarantee that an individual will get AD. Individual’s genetic profile only accounts for a portion of the AD progression risk. In fact, prior work has shown other phenotypes, such as diet, education, environment, and contribute to AD development (Imtiaz et al., 2014). For example, the chance of acquiring AD can be decreased by engaging in regular exercise, eating a balanced diet, and abstaining from tobacco use and excessive alcohol use. Although cerebrospinal fluid (CSF) and positron emission tomography (PET) are valuable for assessing risk factors in Alzheimer’s disease, they are directly or indirectly invasive in nature thereby posing potential health risks (Hansson et al., 2019). Therefore, examining functional and structural brain patterns can offer further insights of brain abnormality, potentially leading to new avenues for treatment strategies. This approach balances the need for precise diagnosis with considerations for patient safety and comfort. All of these mentioned factors can potentially affect brain features as well. Therefore, developing a new AD risk based on brain imaging phenotypes is a promising step in identifying different subgroups of AD.

Magnetic resonance imaging (MRI) technology allows us to collect functional (fMRI) and structural (sMRI) information about the brain noninvasively (Grover et al., 2015). MRI has frequently been used to study structural and functional brain alteration linked to AD (Cuingnet et al., 2011; Jack et al., 2008; Sendi et al., 2023, 2021). Since these functional and structural alterations in the brain might manifest years before symptoms show up, MRI has been studied as a possible technique for the early detection and diagnosis of AD (Abrol et al., 2020; Sendi et al., 2021; Tondelli et al., 2012). MRI can also potentially be used to track the effectiveness of therapy over time and to keep track of how AD develops. However, the majority of studies in the field of AD have primarily focused on case-control comparisons, which offer valuable insights but may not fully capture the individual variability. To achieve a more comprehensive understanding, it is essential to shift our focus towards studying individuals who have not yet received an AD diagnosis. Unfortunately, only a limited number of studies have explored the relationship between clinical indicators of AD and age-related changes in the brain within non-clinical populations. Expanding research in this area would provide valuable insights into early markers, risk factors, and potential interventions for AD in individuals without an existing clinical diagnosis.

This study introduces a novel approach to assess the risk of AD by developing a pipeline for calculating a brain-wide risk score (BRS) using multimodal neuroimaging data. The longitudinal Open Access Series of Imaging Studies (OASIS)-3 cohort served as the reference dataset, enabling the characterization of healthy control (CN) individuals and those with mild cognitive impairment (MCI). Subsequently, the UK Biobank dataset, comprising a substantial sample size (N>37,000), was employed as the target dataset on which we would calculate individual BRS. By leveraging the BRS, we stratified individuals into 10 risk deciles using resting-state functional MRI (rs-fMRI) and 10 risk deciles using structural MRI (sMRI). Additionally, to evaluate the multimodal risk, we stratified participants into two distinct subgroups. These subgroups were focused on the two extreme corners of a spectrum consisting of 100 decile combinations. To further validate our method and the newly developed MCI risk assessment, we conducted additional analyses using the Alzheimer’s Disease Neuroimaging Initiative (ADNI) dataset to confirm similar rs-fMRI and sMRI risk patterns were present in an independent MCI vs control sample. Finally, in the UK Biobank study, we analyzed 46 AD patients and found that both FNC and GM patterns were consistent with those at a higher risk for MCI. We checked for similarities to establish this correlation. These findings emphasize the effectiveness of FNC and sMRI-based BRS in the early detection of AD, suggesting their potential in identifying individuals at an elevated risk for MCI.

## 2. Materials and Methods

### 2.1. Datasets and study population

This study uses two datasets including the OASIS-3 (LaMontagne et al., 2019) cohort as the reference population and UK Biobank (Littlejohns et al., 2020) as the target one. OASIS-3 contains 1389 imaging samples (age: 67.18±8.71) and their associated demographic and clinical data. We used Clinical Dementia Rating Scale Sum of Boxes Scores or CDR_SOB to identify samples with mild cognitive impairment or MCI with CDR_SOB>0 from control or CN with CDR_SOB=0. Overall, we have 1028 sample of CN and 361 sample of MCI in this dataset. The UK Biobank data includes resting-state fMRI (duration: 5min) and sMRI data of 37,780 (20,157 females) adults’ brains and demographic information (age:64.06± 7.51). We additionally used the ADNI dataset (age:71.85± 6.99) which contain 382 CN and 347 MCI individuals to validate our proposed MCI risk score (Mueller et al., 2008; Weber et al., 2021).

### 2.2. Imaging protocol

OASIS-3 data were collected from 3 different Siemens scanners (Siemens Medical Solutions USA, Inc), including one Vision 1.5T and two scanners of TIM Trio 3T with a 16-channel head coil on 1.5T scanners and 20-channel head coil on 3T scanners with foam pad stabilizers placed next to the ears to decrease motion. High-resolution T2*-weighted images were acquired using a gradient-echo EP sequence with TE =27 ms, TR = 2.5 s, flip angle = 90°, slice thickness = 4mm, slice gap = 4 mm, and matrix size = 64 for the Trio scanner. For the Vision scanner, the scanning parameters with gradient-echo EP sequence are TE= 27 ms, TR= 2.2 s, flip angle= 90°, slice thickness=4 mm, slice gap = 4 mm, and matrix size = 64 (LaMontagne et al., 2019).

The UKBiobank imaging data were collected using a 3T Siemens (Siemens Healthineers, Erlangen, Germany) scanner with a 32-channel head coil. T1-weighted structural imaging protocol includes resolution=1×1×1 mm^3^, Duration=4:54 mins, TI/TR=880/2000ms, Field-of-view: 256×256×208. The resting-state fMRI were collected using gradient-echo sequence with resolution=2.4×2.4×2.4 mm^3^, duration=6:10 mins, TE/TR=39/735 ms, field-of-view: =88×88×64 (Miller et al., 2016). The ADNI dataset comprises imaging data acquired using various MRI scanners, including 1.5T GE, 3T GE, 1.5T Philips, 3 T Philips, 1.5T Siemens, and 3T Siemens scanners. The specific details of the imaging protocols utilized in the acquisition of the ADNI dataset can be found in (Song et al., 2022).

### 2.3. Preprocessing and feature extraction

For the resting-state fMRI analysis, the initial step involved removing the first five dummy scans prior to preprocessing. For the subsequent steps, we utilized the default slice timing routines of Statistical Parametric Mapping (SPM12, https://www.fil.ion.ucl.ac.uk/spm/). The slice that was obtained in the middle of the sequence was used as the reference slice in this procedure. For participant head movement correction, we used rigid body motion correction. After that, we used the echo-planar imaging (EPI) template to normalize the imaging data to the standard Montreal Neurological Institute (MNI) space. Finally, we applied a Gaussian kernel with a full width at half maximum (FWHM) of 6mm to smooth the images.

We then used the NeuroMark pipeline to identify a set of independent components for the whole brain of each subject. The NeuroMark_fMRI_1.0 template (available in GIFT; http://trendscenter.org/software/gift; and also at http://trendscenter.org/data) was derived by performing a group independent component analysis (ICA) on two healthy control datasets including the human connectome project (HCP: https://www.humanconnectome.org/study/hcp-young-adult/document/1200-subjects-data-release, 823 subjects after the subject selection) and the genomics superstruct project (GSP: https://dataverse.harvard.edu/dataverse/GSP, 1005 subjects after the subject selection). The extracted ICs from the two datasets were then matched to identify replicable group-level spatial maps. We further evaluated the reproducible ICs pairs by observing their spatial activations and low-frequency fluctuations of their related time courses (TCs). This process yielded 53 ICs and group them into seven domains including subcortical network (SCN), auditory network (ADN), sensorimotor network (SMN), visual network (VSN), cognitive control network (CCN), default-mode network (DMN), and cerebellar network (CBN) (Du et al., 2020). Next, we used this template to perform fully automated spatially constrained ICA, using the GIFT toolbox, to extract component maps and time courses for each subject in the OASIS, ADNI, and UK Biobank data sets. Finally, we implemented additional post-processing steps on the subject-specific time courses to eliminate noise and enhance data quality. These steps included detrending to remove linear, quadratic, and cubic trends, multiple regression of the 6 realignment parameters and their derivatives, identification and removal of outliers, and applying a low-pass filter with a cutoff frequency of 0.15 Hz. It is worth noting that the filtering was specifically performed on the time courses of ICs rather than voxel-based fMRI data, as we aimed to retain more information on fMRI for the subsequent ICA decomposition. This post-processing approach has been successfully utilized in previous studies on ICA-based neuroimaging studies (Sendi et al., 2023, 2021).

We next calculated the functional network connectivity (FNC) for any pairs of ICs of the whole brain using the Pearson correlation between pairs of ICs in each subject as shown in Equation 1:

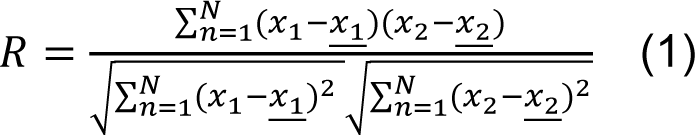

where 𝑥1 and 𝑥2 are time course signals and <u>𝑥1</u> and <u>𝑥2</u> are the mean of 𝑥1 and 𝑥2 , respectively. It takes values in the interval [-1,1] and measures the strength of the linear relationship between 𝑥1 and 𝑥2. Each FNC is a 53×53 symmetric matrix with 53 ICs. Thus, we calculated 1378 connectivity features for each sample.

For the T1-weighted image preprocessing, we first performed spatial registration to a reference brain template in Montreal Neurological Institute (MNI) space. We employed the unified segmentation method integrated within SPM12 software package (https://www.fil.ion.ucl.ac.uk/spm/) to perform the segmentation of T1-weighted images into distinct components, including gray matter, white matter, and cerebrospinal fluid (CSF) images. Unified segmentation utilizes a probabilistic model that combines information from both the T1 - weighted image itself and a priori knowledge about tissue types to estimate the probability of each voxel belonging to gray matter, white matter, or CSF (Ashburner and Friston, 2005). By leveraging this approach, we were able to accurately delineate and separate these tissue types within the T1-weighted images.

### 2.4. Brian-wide risk score estimation

Our proposed approach for neuroimaging based BRS to estimate risk for MCI, shown in Fig.1, includes multiple steps. In the first step (**Generation of references: OASIS-3**), we calculated the mean neuroimaging features across CN and MCI groups in the reference dataset. In the second step (**target population: UK Biobank**), we first calculated the mean of neuroimaging features across all participants and removed it from each participant’s data. Next, we calculated the correlation between the data for each participant in the target dataset and the CN and MCI references generated in step1. Therefore, each target participant has two correlation values, one from each reference group (i.e., R_CN_ and R_MCI_). Finally, we calculated the group difference in the estimated correlations (i.e., Δdiff=R_CN_-R_MCI_). This final value is used as a BRS of MCI in the rest of this paper. We performed the same procedure using the ADNI dataset to validate our method, ensuring the generalizability and reliability of our approach when using different datasets to compute the references.

**Fig.1:**
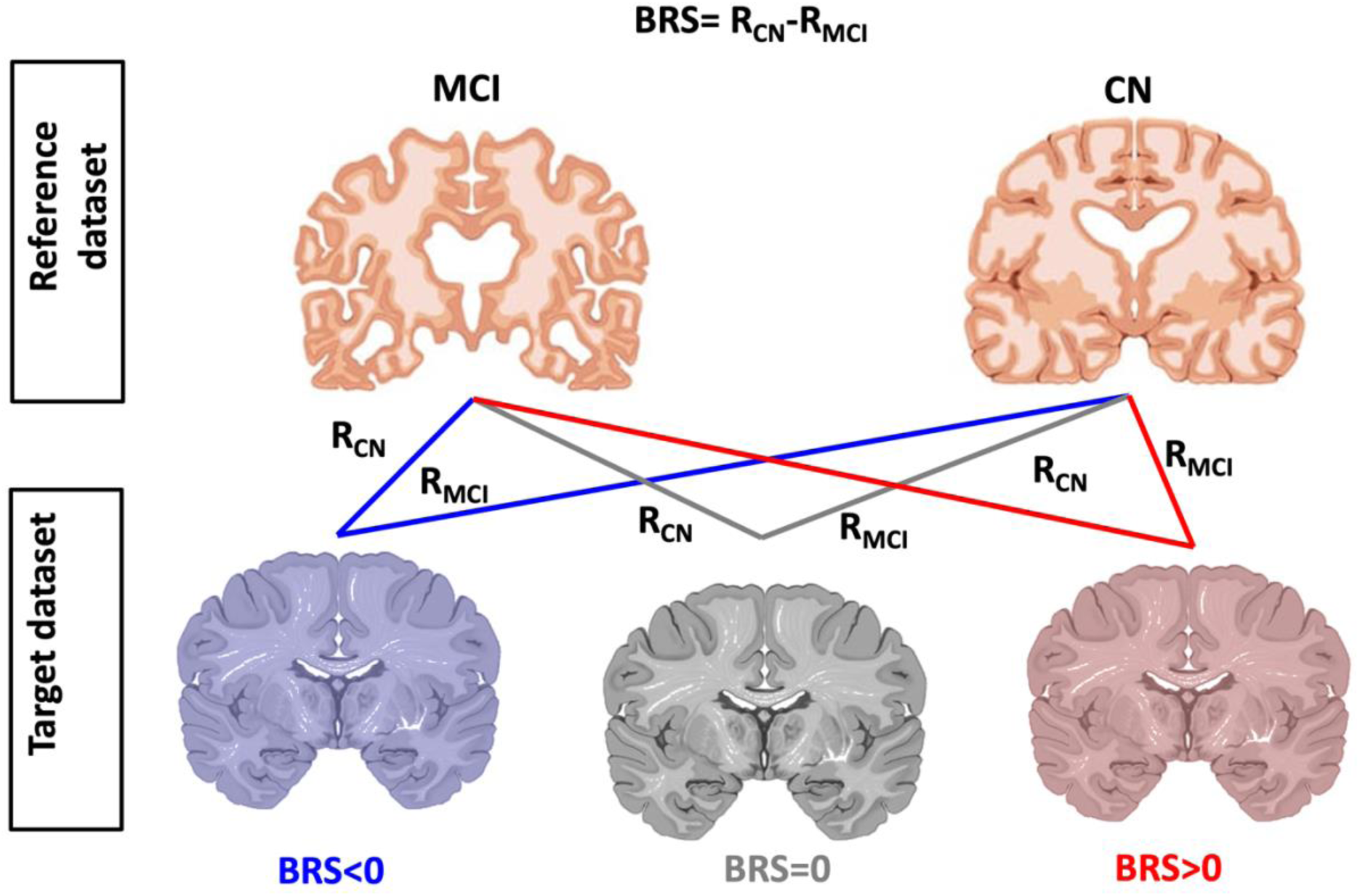
The proposed method for calculating brain-wide risk score (BRS). To begin, we compute the correlation between the brain features of each participant in the UK Biobank dataset with two groups: control or CN (referred to as R_CN_), and mild cognitive impairment or MCI (referred to as R_MCI_), taken from the reference group. Subsequently, we calculate the BRS as the difference between R_CN_ and R_MCI_.

Additionally, to validate the method, we conducted an assessment of the BRS among participants from the OASIS-3 study. The within-sample MCI and CN groups were used as references for this evaluation. Subsequently, we performed a comparative analysis of the BRS between MCI participants and CN participants. Furthermore, we extended our investigation by calculating the BRS of ADNI participants, utilizing the OASIS-3 dataset as a reference for the CN and MCI groups, thus evaluating the BRS in an independent sample. We computed the MCI BRS by employing two methods: one based on FNC derived from resting-state fMRI, and the other using gray matter (GM) estimations obtained from sMRI. Additionally, we similarly evaluated the 46 participants from the UKBB who had been diagnosed with AD.

## 3. Results

### 3.1 Functional network connectivity references for MCI versus controls

In Fig. 2, the mean FNC of the CN (left panel), MCI (middle panel), and CN-MCI (right panel) in the reference dataset (OASIS-3) is displayed. We observed an increased connectivity in sensory networks, including VSN, SMN, and ADN, in the CN group. Functional network connectivity was highly negative in these brain networks for the MCI group. On the other hand, the MCI group showed higher FNC between sensory networks and other brain networks. Specifically, the connectivity between these networks and CBN was highly positive in the MCI group. Furthermore, the CN group exhibited higher SCN and DMN connectivity, whereas this connectivity was less prominent in the brain networks of the MCI group. The second reference dataset, ADNI, displayed a similar FNC pattern in the CN and MCI groups, as shown in Fig. 2B. We conducted a correlation analysis between the CN-MCI differences in both the OASIS and ADNI datasets. This was done to evaluate the similarities between CN-MCI brain features across two datasets. The correlation between the CN-MCI groups in both OASIS-3 (Fig.2A, right panel) and ADNI (Fig.2B, right panel) datasets was found to be 0.67 (N=1378, p<10^-5^).

**Fig.2:**
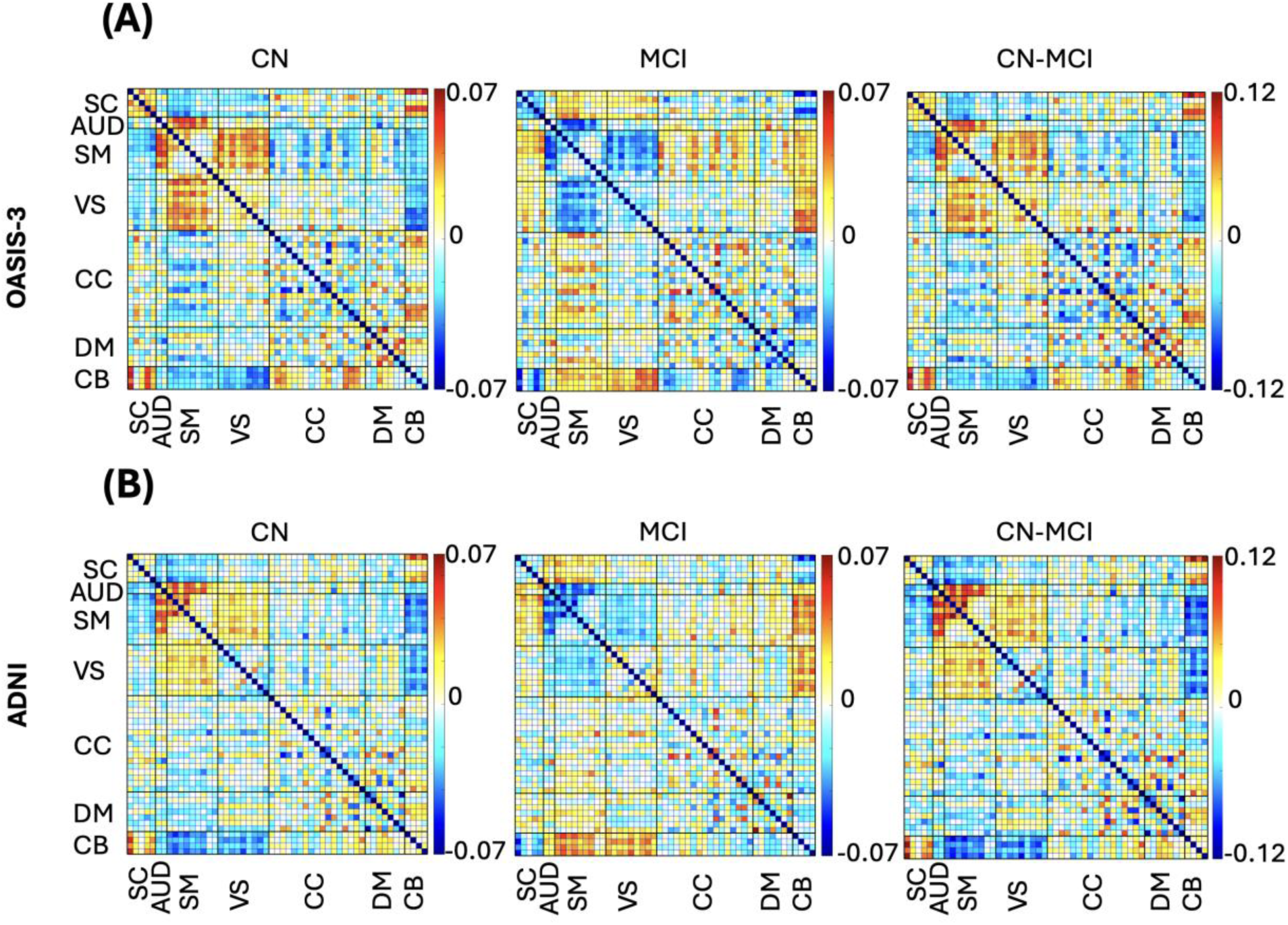
Mean functional network connectivity (FNC) of reference group. **A)** The mean FNC of CN (left), MCI (middle), and CN-MCI (right) in OASIS-3 dataset. **B)** The mean FNC of CN (left), MCI (middle), and CN-MCI (right) in ADNI dataset. The color bar shows the strength of the connectivity. SCN, Subcortical network; ADN, auditory network; SMN, sensorimotor network; VSN, visual network; CCN, cognitive control network; DMN, default-mode network; and CBN, cerebellar network.

### 3.2 Gray matter (GM) map references for MCI versus controls

Fig. 3A presents the GM maps of the CN group (left panel), MCI group (middle panel), and the CN-MCI comparison (right panel) based on the OASIS-3 dataset. As depicted in the figure, we observed higher GM values in the visual, auditory, and sensory-motor networks in the CN group. This finding aligns with our previous observations of increased sensory network connectivity in the CN group FNC. Furthermore, the CN group also exhibited higher GM values in the cerebellar regions. Interestingly, we observed similar patterns in the GM comparison between the CN and MCI groups in the ADNI dataset, as we did in the OASIS-3 dataset. Specifically, higher GM values were found in the visual, auditory, and sensory-motor networks in the CN group when compared to the MCI group.

**Fig.3:**
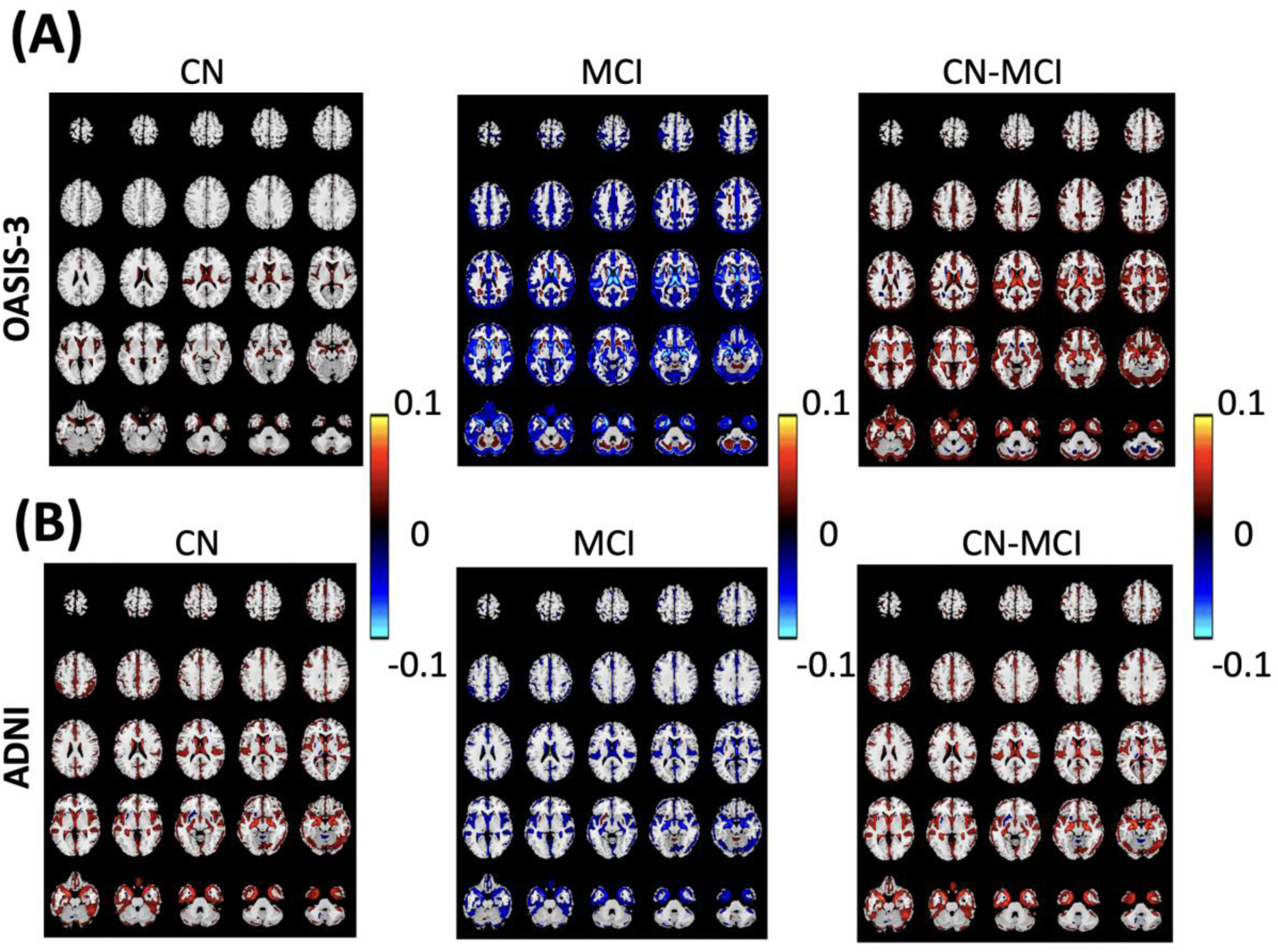
Mean gray matter (GM) of reference group. **A)** The mean GM of CN (left), MCI (middle), and CN-MCI (right) in OASIS-3 dataset. **B)** The mean GM of CN (left), MCI (middle), and CN-MCI (right) in ADNI dataset. The color bar shows the strength of the gray matter.

### 3.3 Validation of the BRS in OASIS-3 and ADNI dataset

In this study, we employed the MCI and CN groups from the OASIS-3 dataset as reference cohorts and calculated the BRS for the participants within this dataset. Fig. 4A presents a 2D histogram of the BRSCN (left panel), BRSMCI (middle panel), and BRSCN-BRSMCI (right panel) groups of OASIS-3 dataset. The figure demonstrates the clear differentiation between the MCI and CN groups based on the BRS we calculated for the OASIS-3 dataset. The average FNC BRS for the CN group is 0.0361, whereas for the MCI group, it is -0.1221. Furthermore, considering the sMRI BRS, the CN group has an average score of 0.0504, while the MCI group has a score of -0.2091. To further validate our results, we employed a five-fold cross-validation approach. In this process, we divided the OASIS-3 dataset into 80% reference data and 20% target data segments. We then calculated the BRS for the target group and compared it with the BRS of the CN and MCI groups within the target segment. This procedure was repeated five times, ensuring coverage of the entire OASIS-3 dataset. The obtained p-values, using analysis of variance or ANOVA, for FNC BRS ranged from 2.1296 x10^-18^ (minimum) to 9.9197 x10^-14^ (maximum), while the p-values for sMRI BRS ranged from 3.4210 x10^-35^ (minimum) to 2.9487 x10^-29^ (maximum). Using 2D BRS (i.e., considering both FNC BRS and sMRI BRS together), the obtained p-values, from Multivariate analysis of variance or MANOVA, raged from 2.9445 x10^-18^ (minimum) to 4.1647 x10^-14^ (maximum).

**Fig.4:**
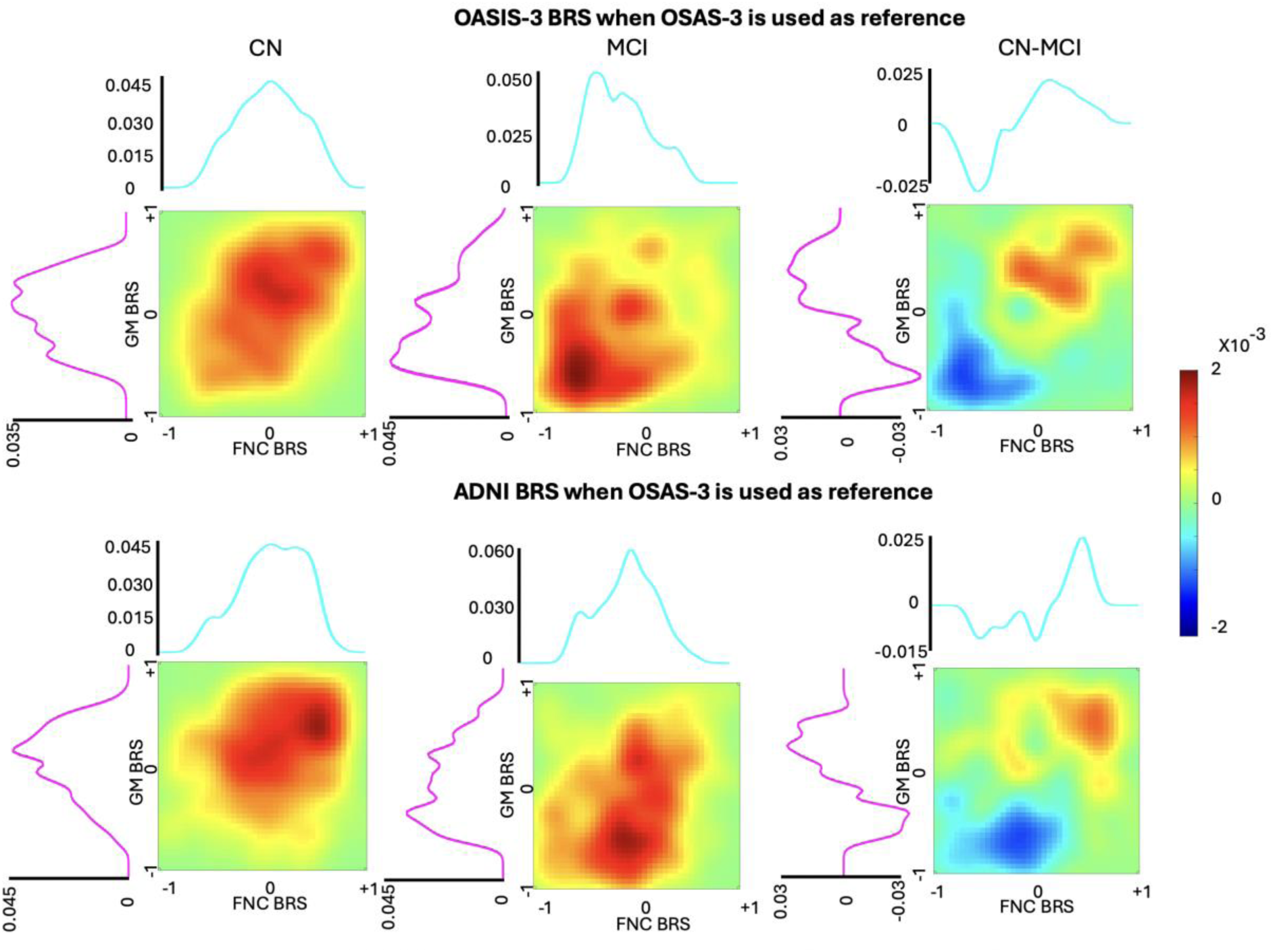
Validating BRS in OASIS-3 and ADNI. **A)** 2D histogram of BRS_CN_, BRS_MCI_, and BRS_CN_- BRS_MCI_ in OASIS-3 dataset in which we used OASIS-3 as the reference group. **B)** 2D histogram of BRS_CN_, BRS_MCI_, and BRS_CN_-BRS_MCI_ in ADNI dataset in which we used OASIS-3 as the reference group.

Fig. 4B displays the 2D histogram plot of BRS_CN_ (left panel), BRS_MCI_ (middle panel), and BRS_CN_- BRS_MCI_ (right panel) groups in the ADNI dataset. For this analysis, we utilized OASIS-3 as the reference dataset, thus it was completely independent of the ADNI dataset. Similar to Fig. 4A, our observations demonstrate a distinct separation of BRS between the MCI and CN groups, further affirming the accuracy of the BRS method in effectively distinguishing between these two populations. The average FNC BRS for the CN (i.e., BRS_CN_) and MCI (i.e., BRS_MCI_) groups are 0.0583 and -0.0297, respectively. An ANOVA test reveals a statistically significant difference between these means (p = 7.1703 x10^-7^). Moreover, considering the sMRI BRS, the BRS_CN_ group has an average score of 0.0855, while the BRS_MCI_ group has a score of -0.0869 (p =2.7572 x10^-13^). Additionally, when employing 2D BRS, which encompasses both FNC BRS and sMRI BRS, the resulting p-value was found to be 1.9678 x10^-14^.

### 3.4 Identified MCI subgroups based on FNC risk deciles

After calculating the BRS of participants in the UKBB based on each reference dataset, we divided the scores into deciles. We then categorized each participant based on their respective risk decile, resulting in the identification of 10 subgroups. In Fig. 5A, we present the 10 subgroups that were identified using the OASIS-3 dataset as a reference. To assess the similarity between each subgroup and the MCI and CN groups, we measured the correlation between the mean FNC of each subgroup and the mean FNC of each group, as depicted in Fig. 5B. Each subgroup represents a unique grouping of participants based on their BRS. Notably, subgroup 1 closely resembles the characteristics of the MCI group, suggesting that individuals assigned to subgroup 1 exhibit BRS patterns similar to those typically observed in individuals with MCI. Conversely, subgroup 10 displays similarities with the CN group, indicating that participants assigned to subgroup 10 possess BRS patterns that closely align with individuals who have normal cognitive function. Based on Fig. 5B, we can identify subgroups 7, 8, 9, and 10 as CN, and subgroups 1, 2, 3, 4, 5, and 6 as MCI. Additionally, Fig. 5C showcases the mean FNC of each subgroup. It is evident that the mean FNC of subgroup 1 is similar to the mean FNC of the MCI group, while the mean FNC of subgroup 10 is similar to the mean FNC of the CN group.

**Fig. 5:**
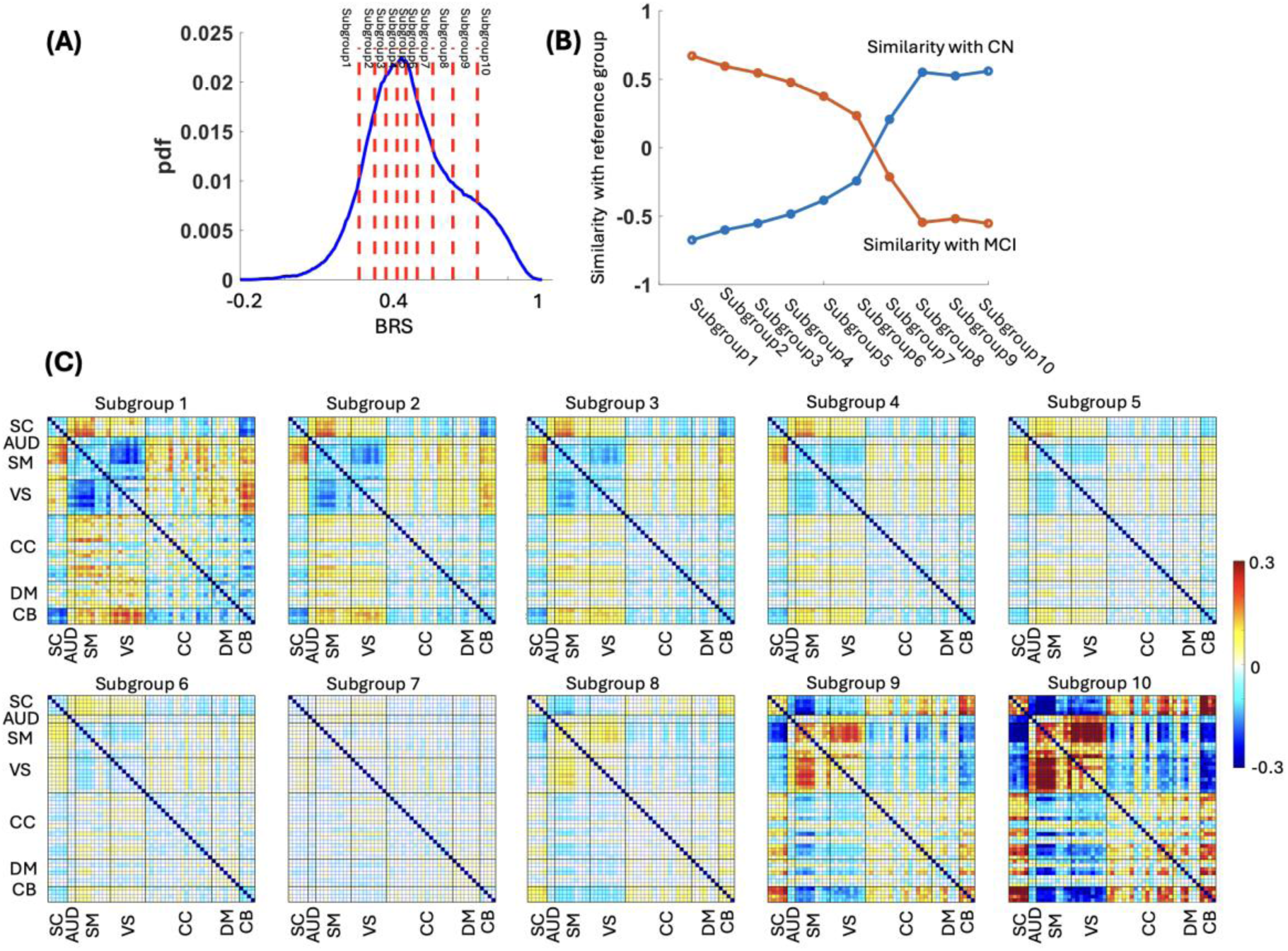
Subgroups of mild cognitive impairment in UKBB based on FNC BRS with OASIS-3 as reference group. **A)** Ten subgroups were identified based on the decile of Brain-wide Risk Score (BRS) Functional Network Connectivity (FNC) in the OASIS-3 dataset. **B)** The correlation between the average FNC of each subgroup and the reference groups of Mild Cognitive Impairment (MCI) and Control (CN). Based on this graph, subgroup 1 resembles the MCI group, while subgroup 10 resembles the CN group. **C)** The average FNC of each subgroup identified based on FNC BRS.

The results were replicated using the ADNI dataset, as illustrated in Fig. 6A. We calculated the correlation between the mean FNC of each subgroup and the mean FNC of the CN and MCI groups, as shown in Fig. 6B. Similar to the findings in Fig. 5B, subgroups 1, 2, 3, 4, 5, and 6 exhibited similarities to the MCI group of the reference dataset, while the remaining subgroups were more similar to the CN group. The replication of our findings using the ADNI dataset as a reference confirmed the robustness and consistency of our results. The identified subgroups exhibited similarities to the MCI and CN groups, supporting the notion that these subgroups capture meaningful patterns in BRS.

**Fig. 6:**
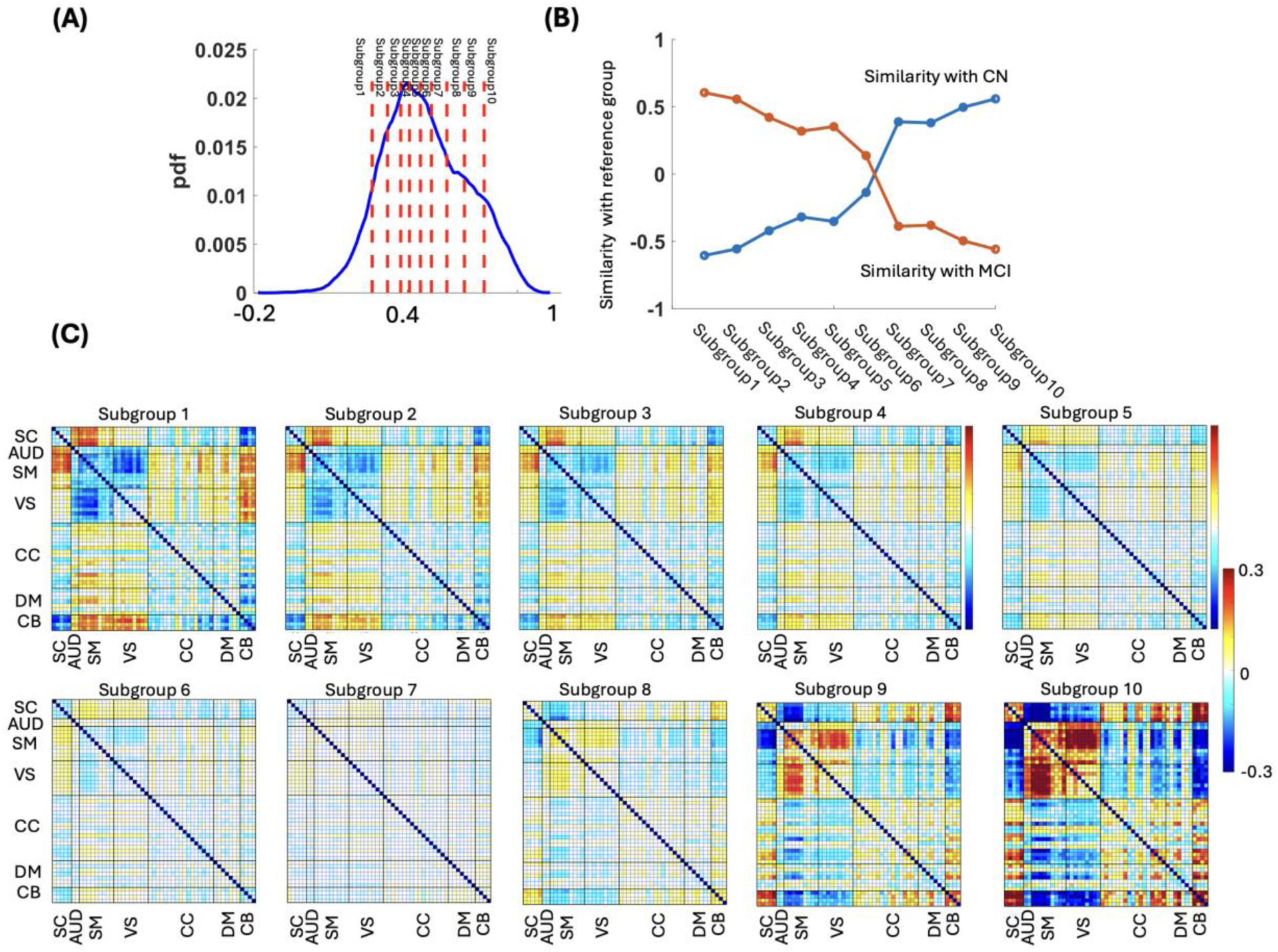
Subgroups of mild cognitive impairment in UKBB based on FNC BRS with ADNI as reference group. **A)** Ten subgroups were identified based on the decile of Brain-wide Risk Score (BRS) Functional Network Connectivity (FNC) in the ADNI dataset. **B)** The correlation between the average FNC of each subgroup and the reference groups of Mild Cognitive Impairment (MCI) and Control (CN). Based on this graph, subgroup 1 resembles the MCI group, while subgroup 10 resembles the CN group. **C)** The average FNC of each subgroup identified based on FNC BRS.

### 3.5 Identified MCI subgroups based on GM risk deciles

Similar to the previous section, we categorized the participants in the UK Biobank dataset into 10 distinct subgroups based on their respective GM-based BRS deciles, as illustrated in Fig. 7A. Additionally, we computed the correlation between the mean GM of each subgroup and the mean GM of the CN and MCI groups, as depicted in Fig. 7B. Based on these correlation results, we identified subgroups 1, 2, 3, 4, and 5 as resembling the MCI group, while subgroups 6, 7, 8, 9, and 10 exhibited similarities to the CN group. Furthermore, Fig. 7C displays the mean GM of each subgroup, confirming the same pattern. To enhance clarity and visualization, we focused on presenting the data from two specific subgroups: those with lower BRS, namely Subgroup1-3, and those with higher BRS, identified as Subgroup 8-10. Subgroups 1-3 demonstrate characteristics similar to the MCI group (Fig.3A left panel), while subgroups 8-10 exhibit features comparable to the CN group, aligning with the findings in Fig. 3A (middle panel). Furthermore, we observed similar patterns when using the ADNI dataset to identify the MCI and CN groups, as shown in Fig. 8. In this case, we identified 5 subgroups that resembled the CN group and another 5 subgroups that resembled the MCI group, as depicted in Fig. 8B and 8C, respectively.

**Fig. 7:**
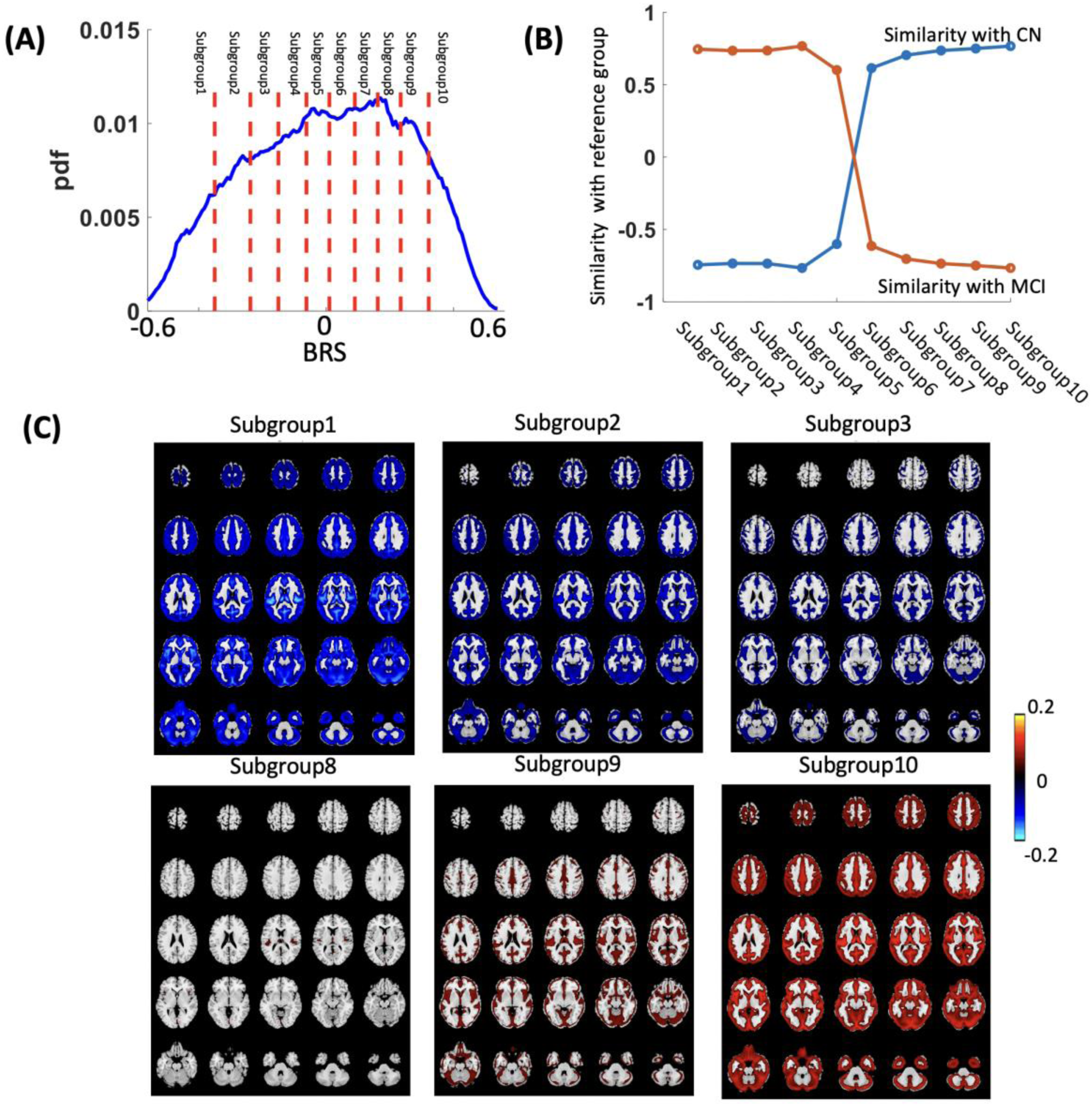
Subgroups of mild cognitive impairment in UKBB based on GM BRS with OASIS-3 as reference group. **A)** Ten subgroups were identified based on the decile of Brain-wide Risk Score (BRS) gray matter (GM) in the OASIS-3 dataset. **B)** The correlation between the average GM of each subgroup and the reference groups of Mild Cognitive Impairment (MCI) and Control (CN). Based on this graph, subgroup 1 resembles the MCI group, while subgroup 10 resembles the CN group. **C)** The average GM of each subgroup identified based on GM BRS. For clearer understanding and better visualization, our presentation is tailored to showcase data specifically from two contrasting subgroups: the first consisting of Subgroup1-3, characterized by lower BRS, and the second comprising Subgroup 8-10, noted for their higher BRS.

**Fig. 8:**
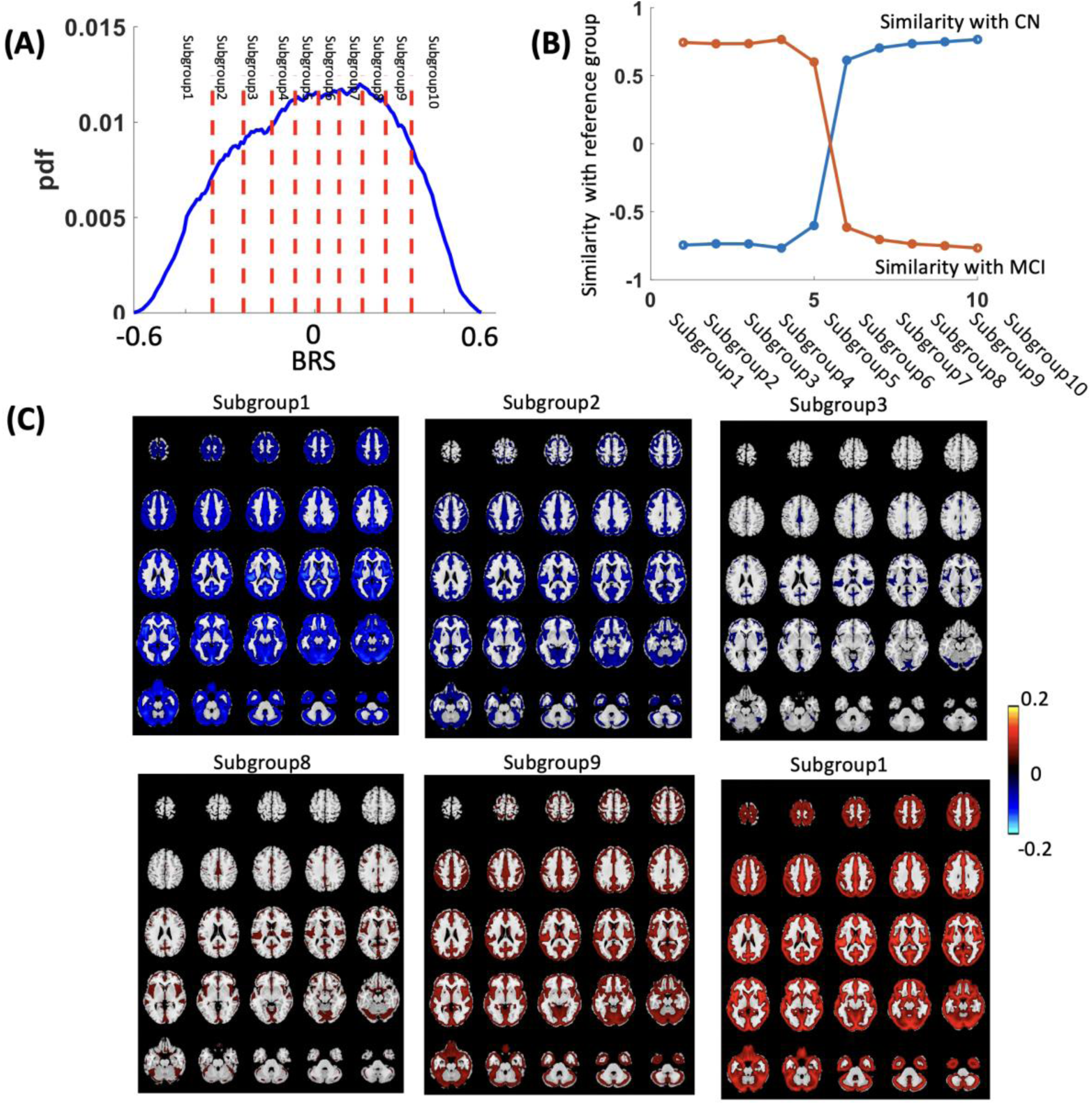
Subgroups of mild cognitive impairment in UKBB based on GM BRS with ANDI as reference group. **A)** Ten subgroups were identified based on the decile of Brain-wide Risk Score (BRS) gray matter (GM) in the ADNI dataset. **B)** The correlation between the average GM of each subgroup and the reference groups of Mild Cognitive Impairment (MCI) and Control (CN). Based on this graph, subgroup 1 resembles the MCI group, while subgroup 10 resembles the CN group. **C)** The average GM of each subgroup identified based on GM BRS. For clearer understanding and better visualization, our presentation is tailored to showcase data specifically from two contrasting subgroups: the first consisting of Subgroup1-3, characterized by lower BRS, and the second comprising Subgroup 8-10, noted for their higher BRS.

### 3.6 Identified MCI subgroups based on multimodal risk

By combining 10 deciles each from FNC-based BRS and GM-based BRS, we derived 100 distinct deciles. From these, we categorized participants into two groups based on their risk of MCI. The low-risk group consists of participants who are in both the 1^st^ decile of FNC-based BRS and the 1^st^ decile of GM-based BRS. Conversely, the high-risk group includes participants who are in both the 10^th^ decile of FNC-based BRS and the 10^th^ decile of GM-based BRS. Fig.9A illustrates the two groups identified using the OASIS-3-based BRS, while Fig. 9B demonstrates the groups as identified through the ADNI-based approach. The left panel of Fig.10A illustrates the similarities between the low-risk and high-risk groups in relation to the CN and MCI reference groups, which are derived from the OASIS-3 dataset. The middle panel of Fig.10A displays the mean FNC of the high-risk group, while the right panel shows the mean FNC of the low-risk group. Analysis of Fig.10A reveals that the FNC of the low-risk group closely resembles that of the CN group, and the FNC of the high-risk group is similar to that of the MCI group. Additionally, the left panel of Figure 10B highlights the similarities in GM between the two groups categorized by MCI risk according to the multimodal BRS and compares these with the GM of the MCI and CN reference groups from OASIS-3. Upon closer examination, it is observed that the GM pattern of the high-risk group (shown in the middle panel of Fig. 10B) closely aligns with that of the MCI group. Similarly, the GM pattern of the low-risk group (depicted in the right panel of Fig. 10B) closely resembles that of the CN group. The results were consistently replicated using ADNI samples as the reference group, as depicted in Figure 11. Specifically, Figure 11A demonstrates that the low-risk group, identified through multimodal BRS, exhibits an FNC pattern similar to that of the CN group, while the high-risk group displays an FNC pattern akin to that of the MCI group. Furthermore, as illustrated in Figure 11B, it was observed that the GM pattern of the low-risk group closely resembles that of the CN group, and the GM pattern of the high-risk group mirrors that of the MCI group within the ADNI samples.

**Fig. 9:**
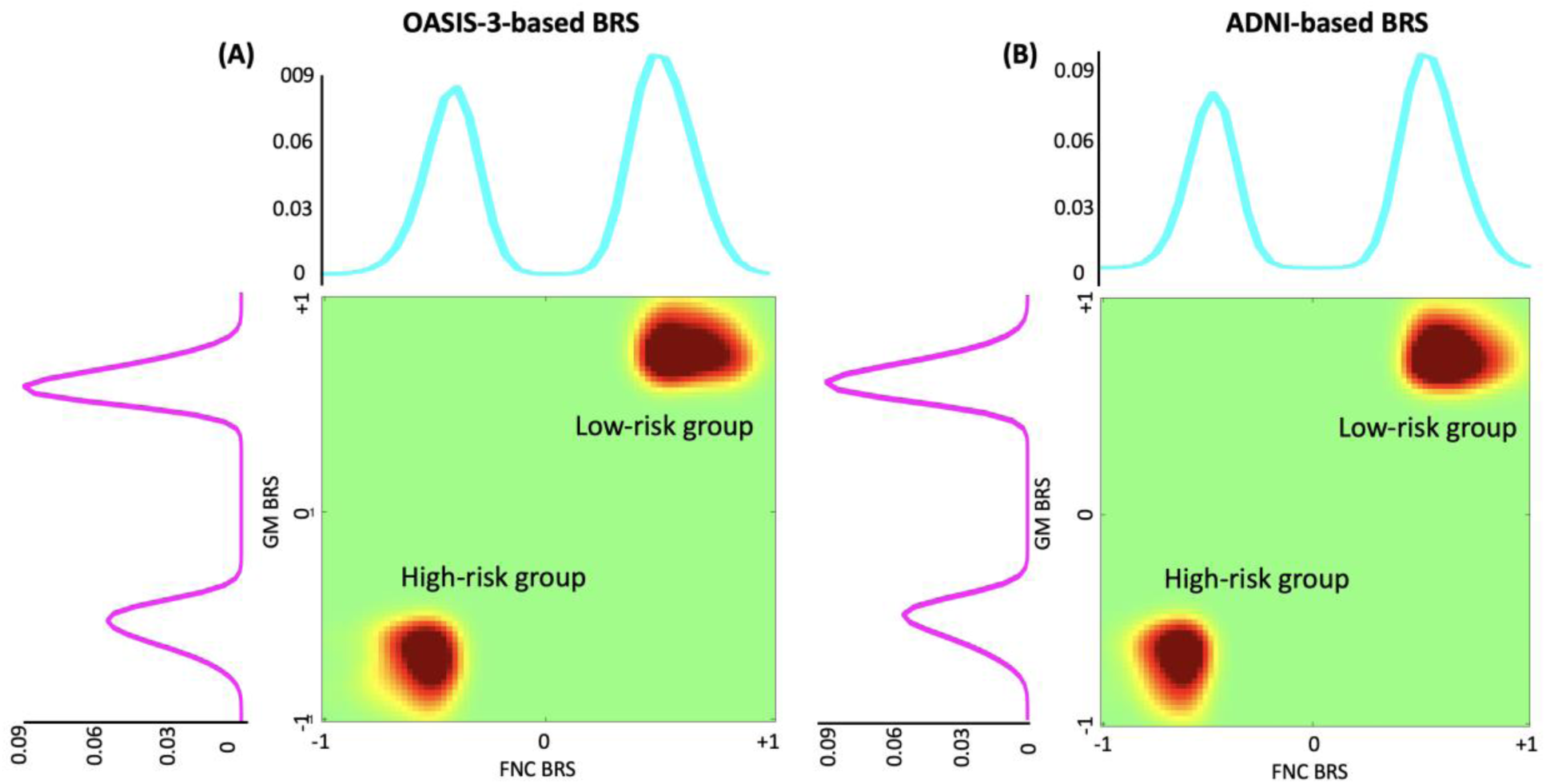
Subgroups of mild cognitive impairment in UKBB based on multimodal BRS. **A)** Using OASIS-3 as the reference group, we identified two distinct groups of UKBB participants categorized by higher and lower risk of Mild Cognitive Impairment (MCI), based on assessments from multimodal BRS. B) Using ADNI as the reference group, we identified two distinct groups of UKBB participants categorized by higher and lower risk of MCI, based on assessments from multimodal BRS.

**Fig. 10:**
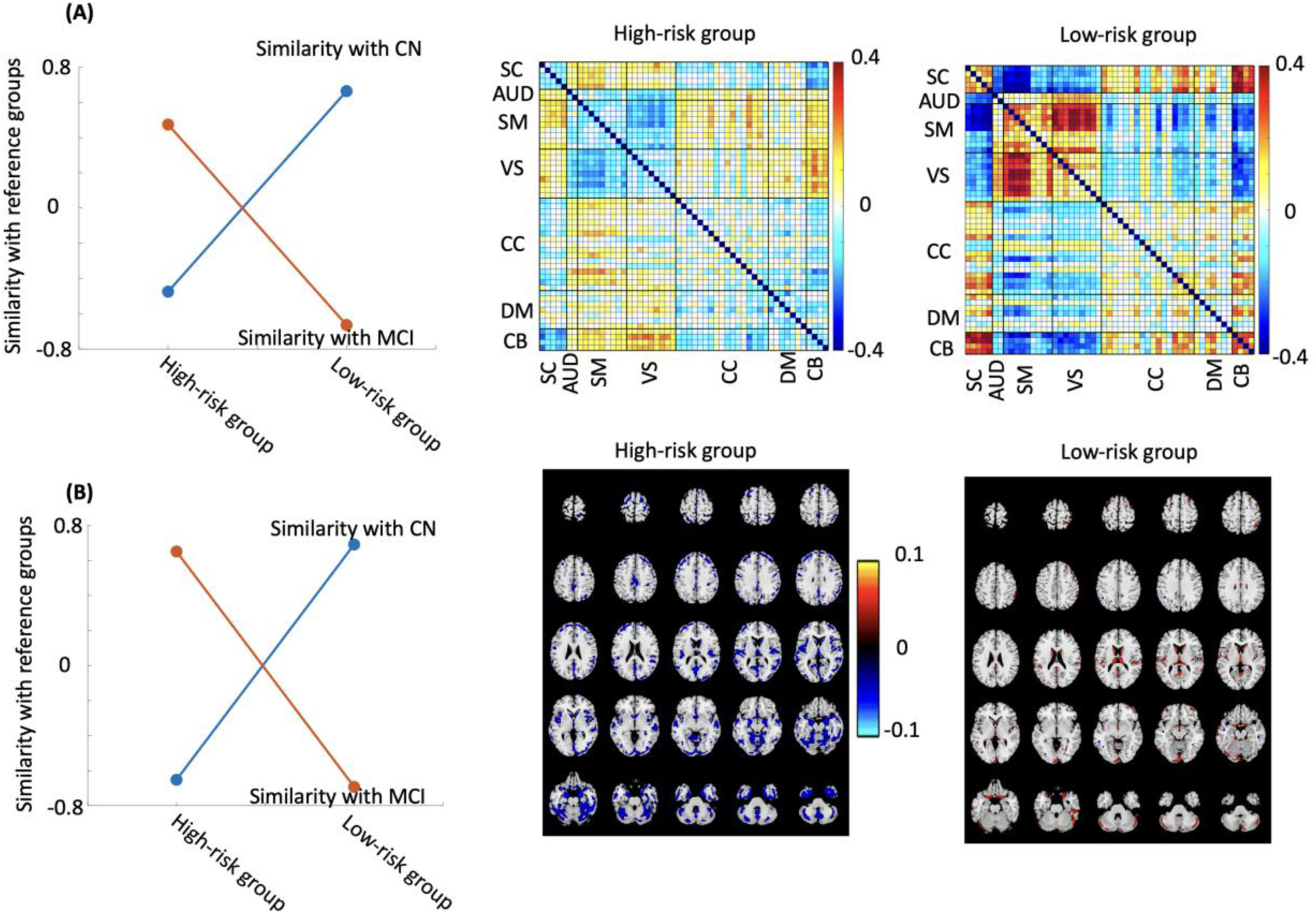
Assessing similarity between participants identified using multimodal BRS and the CN and MCI reference groups from OASIS-3. **A)** This part demonstrates the similarity in Functional Network Connectivity (FNC) between groups with high and low MCI risk compared to the reference CN and MCI groups**. B)** This part demonstrates the similarity in gray matter (GM) between groups with high and low MCI risk compared to the reference CN and MCI groups.

**Fig. 11:**
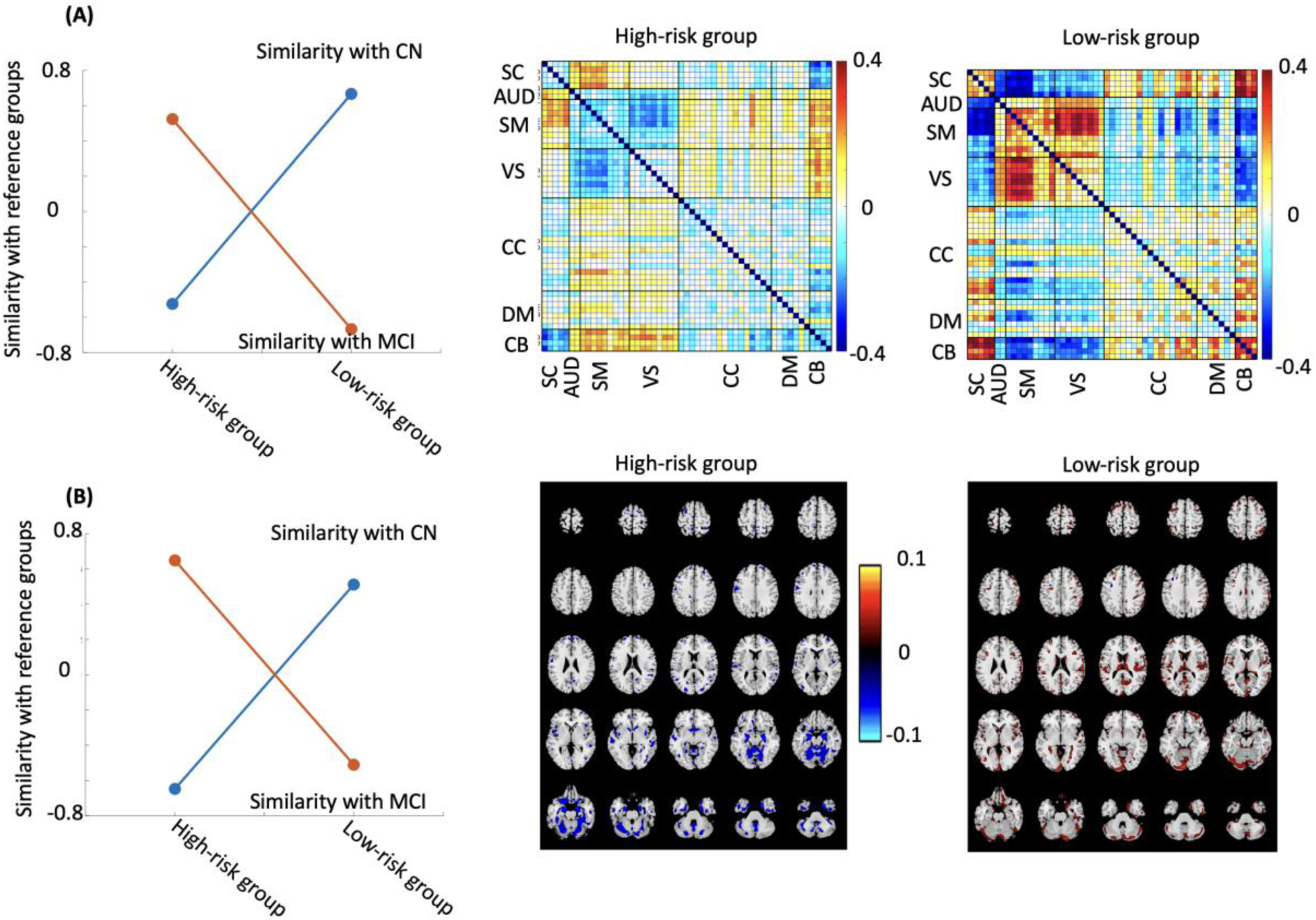
Assessing similarity between participants identified using multimodal BRS and the CN and MCI reference groups from ADNI. **A)** This part demonstrates the similarity in Functional Network Connectivity (FNC) between groups with high and low MCI risk compared to the reference CN and MCI groups**. B)** This part demonstrates the similarity in gray matter (GM) between groups with high and low MCI risk compared to the reference CN and MCI groups.

### 3.7 Efficacy of BRS in early AD detection

We identified 46 UKBB participants diagnosed with AD within 5 years following their neuroimaging data collection. We then determined the number of AD-diagnosed participants across all 10 subgroups, as classified using FNC-and sMRI-based BRS. Fig.12A and Fig.12B present the mean FNC and mean GM, respectively, of the 46 UKBB participants. Additionally, Fig.12C illustrates the similarities between the mean FNC and GM of these UKBB participants and those of a subgroup identified using the BRS decile. It was observed that UKBB participants diagnosed with AD exhibit FNC and GM patterns that are similar to the subgroup at a higher risk of MCI. This result was consistently replicated using the ADNI as the reference group, as shown in Fig.12D. The results demonstrated consistency with the use of multimodal BRS, as illustrated in Fig. 12E (using OSAIS-3 as the reference group) and Fig. 12F (using ADNI as the reference group). These figures reveal that participants from the UKBB subsequently diagnosed with AD exhibit FNC and GM patterns akin to those observed in the high-risk groups.

**Fig. 12:**
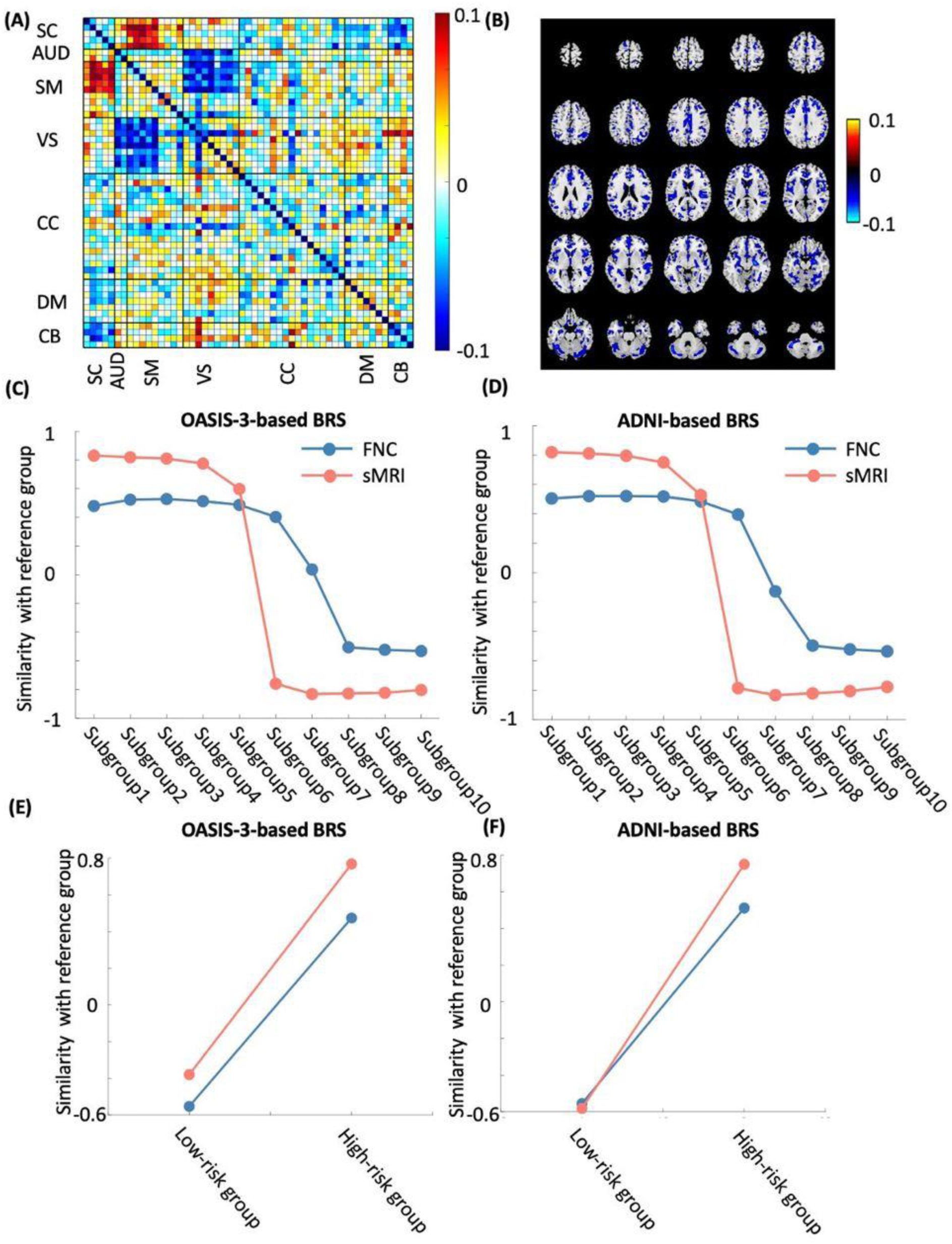
Assessing similarity between UKBB participants diagnosed for AD functional network connectivity (FNC)-based, gray matter (GM)-based, and multimodal brain-wide risk score (BRS). **A)** Mean of FNC of UKBB participants diagnosed for AD. **B)** Mean of GM of UKBB participants diagnosed for AD. **C)** Similarity measure between UKBB participants diagnosed for AD and risk group identified in UKBB when we used OASIS-3 as a reference group. **D)** Similarity measure between UKBB participants diagnosed for AD and risk group identified in UKBB when we used ADNI as a reference group. **E)** Similarity measure between UKBB participants diagnosed for AD and low/high risk group based on multimodal BRS when we used OASIS-3 as a reference. **F)** Similarity measure between UKBB participants diagnosed for AD and low/high risk group based on multimodal BRS when we used ADNI as a reference.

## 4. Discussion

The risk of AD pertains to the probability or likelihood of an individual developing this neurodegenerative disorder (Knopman et al., 2021). Numerous factors, encompassing genetic and environmental influences, contribute to the risk of AD. Nevertheless, prior research has primarily concentrated on investigating the genetic risk associated with AD (Stocker et al., 2018). It is imperative to recognize that the development of AD is influenced by both genetic and environmental factors. Considering this, the present study has introduced a novel approach to assess the risk of AD by leveraging neuroimaging data, which is referred to as the brain risk score (BRS). This new risk score incorporates relevant information from neuroimaging to provide a comprehensive evaluation of an individual’s risk for developing AD.

We utilized two separate datasets, namely OASIS-3 and ADNI, to establish the reference groups for CN and MCI individuals. To define these reference groups, we employed FNC and GM map mapping techniques. By leveraging these datasets and employing FNC and GM mapping, we were able to form robust reference groups for CN and MCI individuals, facilitating a comprehensive analysis of MCI BRS in UK biobank. In our analysis of FNC data, we observed that the FNC patterns differed between the CN and MCI groups. Specifically, we found that the sensory network FNC exhibited greater prominence in the CN group compared to the MCI group. This finding is consistent with previous studies that have highlighted the importance of sensory network connectivity in predicting the progression of AD (Sendi et al., 2023, 2021). Importantly, we were able to replicate these results in an independent dataset (i.e., ADNI), confirming the robustness of our findings. These findings suggest that FNC, particularly in the sensory network, may serve as a valuable biomarker for distinguishing between CN and MCI individuals, providing insights into the potential role of FNC in understanding AD development and progression (Cai et al., 2015; Li et al., 2020). While the current study primarily focused on analyzing whole brain FNC and defining the MCI BRS, future research may benefit from specifically examining the sensory network FNC. It would be valuable to explore whether a sensory network-based BRS could potentially serve as a superior predictor of AD compared to the whole brain-based MCI BRS.

Through our analysis of the GM map from the OASIS-3 dataset, we found a distinct GM loss distributed across numerous brain regions. This observation is consistent with previous research indicating a combination of global and local GM loss in individuals diagnosed with MCI and AD when compared to the CN group (Karas et al., 2004). The identified GM loss across various brain regions further supports the notion of a widespread and potentially progressive degenerative process in individuals with MCI and AD. Notably, we found a more pronounced reduction in the GM of the visual sensory, auditory, and sensorimotor networks, highlighting the significant involvement of these specific brains and their particular susceptibility to the pathological changes associated with AD.

In our research, we broadened our scope beyond the standard neuroimaging analysis of the CN and MCI groups found in our reference datasets. Our objective is to predict MCI risk within the UKBB population, using OASIS-3 and ADNI as our reference datasets. Diverging from the conventional approach of focusing solely on genetic factors for MCI risk prediction, we established a novel methodology. Our approach subtly incorporates both genetic and environmental factors influencing MCI risk by utilizing neuroimaging data as a proxy. This integration is possible because the brain data we use is susceptible to both genetic and environmental influences. Our research marks a significant step in MCI risk evaluation within the UKBB dataset, highlighting the crucial role of combining functional and structural MRI data. Our findings reveal a powerful synergy between these two types of MRI data in assessing MCI risk. Notably, our analysis within the UKBB cohort showed that individuals with higher BRS closely resembled the CN group, suggesting a reduced risk of MCI. In contrast, those with lower BRS mirrored the MCI group, indicating a higher MCI risk. Previous studies demonstrated the effectiveness of combining resting-state fMRI functional connectivity analysis and sMRI data (Dang et al., 2023; Hojjati et al., 2019). Using machine learning techniques, to accurately identify early stages of Alzheimer’s disease among various groups, a recent study achieved a maximum classification accuracy of 67% and 56% for different group classification using multimodal neuroimaging technique (Hojjati et al., 2019). What distinguishes our study is the consistency and validation of our findings when replicated with independent datasets, specifically OASIS-3 and ADNI. These datasets are considered benchmarks in the field. This replication not only reaffirms the solidity of our research but also highlights the transformative potential of merging functional and structural MRI data for accurate MCI risk assessment within the UKBB group.

It is important to note that recent research has indicated that AD genetic risk can impact both functional and structural neuroimaging data (Chandler et al., 2022; Cho et al., 2035; Mirza-Davies et al., 2022; Sendi et al., 2023). Building upon this understanding, we hypothesized that our newly developed BRS would be capable of accounting for both genetic and environmental factors. This novel approach to MCI risk estimation offers a more comprehensive perspective on the complex interplay of genetic and environmental factors in the development of the disease. By acknowledging that MCI risk is not solely determined by genetics, but also influenced by environmental factors, this can enhance our understanding of the disease’s multifactorial nature. This knowledge has implications for personalized risk assessment, early detection, and the development of targeted interventions that consider both genetic and environmental risk factors.

Our research highlights a significant synergy in identifying MCI risk, particularly emphasizing the effectiveness of sMRI-based BRS as a biomarker for early detection and classification of AD. This is pivotal since MCI often precedes AD, and early detection is crucial for effective treatment. Our findings demonstrate that both unimodal and multimodal BRS are effective in AD detection. However, we observed that sMRI-based BRS and multimodal-based BRS outperform FNC-based BRS in identifying AD. This distinction underlines the potential of sMRI-based BRS and multimodal-based BRS as more precise and reliable tools in detecting early-stage AD, thereby enhancing the accuracy of AD diagnosis. The implications of our findings are substantial, suggesting that sMRI-based BRS and multimodal-BRS could play a vital role in early and accurate identification of AD.

Our study has several limitations that warrant consideration. Firstly, in this current study, we applied our method to the entire brain dataset, which may have introduced some noise as certain brain regions and features might not effectively distinguish MCI from CN individuals within our reference group. Therefore, future research concentrating on these specific brain regions, aimed at enhancing the discrimination between MCI and CN, holds the potential to improve the accuracy of the BRS for our target population. In the current study we do not compare polygenic risk score (PRS) and BRS, future study is needed to compare at the predictability power of BRS and PRS in predicting AD in UKBB. Furthermore, in our current study, we employed correlation as the distance metric. Future research should aim to investigate the potential advantages and limitations of alternative distance metrics in estimating the BRS.

In conclusion, our analysis of the BRS scores obtained from participants in the UKBB using different reference datasets allowed us to identify 10 distinct subgroups based on FNC BRS, 10 subgroups based on GM BRS, and 2 subgroups based on multimodal BRS. Through the correlation analysis of the mean FNC of each subgroup with the CN and MCI groups, we found that subgroup 1 demonstrated close resemblance to the MCI group, while subgroup 10 exhibited similarities with the CN group. These consistent findings were replicated when using the ADNI dataset as a reference. These results suggest that the identified subgroups capture meaningful patterns in BRS and provide insights into the cognitive status of individuals within the study population. Further investigation of these subgroups may contribute to our understanding of cognitive impairment and potentially aid in the identification and characterization of individuals at risk for developing cognitive disorders.

## 6. CRediT authorship contribution statement

Elaheh Zendehrouh: Conceptualization, Methodology, Data curation, Writing – original draft. Mohammad Sendi: Conceptualization, Methodology, and Writing – original draft. Anees Abrol: Writing – review & editing. Ishaan Batta: Writing – review & editing. Reihaneh Hassanzadeh: Writing – review & editing. Vince D. Calhoun: Conceptualization, Methodology, and Writing – review & editing.

## Data Availability

All data produced in the present study are available upon reasonable request to the authors

## References

1. Abrol, A., Bhattarai, M., Fedorov, A., Du, Y., Plis, S., Calhoun, V., 2020. Deep residual learning for neuroimaging: An application to predict progression to Alzheimer’s disease. J Neurosci Methods 339. 10.1016/j.jneumeth.2020.108701

2. Ashburner, J., Friston, K.J., 2005. Unified segmentation. Neuroimage 26, 839–851. 10.1016/j.neuroimage.2005.02.018

3. Cai, S., Chong, T., Zhang, Y., Li, J., von Deneen, K.M., Ren, J., Dong, M., Huang, L., 2015. Altered functional connectivity of fusiform gyrus in subjects with amnestic mild cognitive impairment: A resting-state fMRI study. Front Hum Neurosci 9. 10.3389/fnhum.2015.00471

4. Chandler, H., Wise, R., Linden, D., Williams, J., Murphy, K., Lancaster, T.M., 2022. Alzheimer’s genetic risk effects on cerebral blood flow across the lifespan are proximal to gene expression. Neurobiol Aging 120, 1–9. 10.1016/j.neurobiolaging.2022.08.001

5. Cho, S., Lee, H., Seo, J., 2021. Impact of Genetic Risk Factors for Alzheimer’s Disease on Brain Glucose Metabolism. 10.1007/s12035-021-02297-x/Published

6. Crous-Bou, M., Minguillón, C., Gramunt, N., Molinuevo, J.L., 2017. Alzheimer’s disease prevention: From risk factors to early intervention. Alzheimers Res Ther. 10.1186/s13195-017-0297-z

7. Cuingnet, R., Gerardin, E., Tessieras, J., Auzias, G., Lehéricy, S., Habert, M.O., Chupin, M., Benali, H., Colliot, O., 2011. Automatic classification of patients with Alzheimer’s disease from structural MRI: A comparison of ten methods using the ADNI database. Neuroimage 56, 766–781. 10.1016/j.neuroimage.2010.06.013

8. Dang, C., Wang, Y., Li, Q., Lu, Y., 2023. Neuroimaging modalities in the detection of Alzheimer’s disease-associated biomarkers. Psychoradiology. 10.1093/psyrad/kkad009

9. Du, Y., Fu, Z., Sui, J., Gao, S., Xing, Y., Lin, D., Salman, M., Abrol, A., Rahaman, M.A., Chen, J., Hong, L.E., Kochunov, P., Osuch, E.A., Calhoun, V.D., 2020. NeuroMark: An automated and adaptive ICA based pipeline to identify reproducible fMRI markers of brain disorders. Neuroimage Clin 28. 10.1016/j.nicl.2020.102375

10. Grover, V.P.B., Tognarelli, J.M., Crossey, M.M.E., Cox, I.J., Taylor-Robinson, S.D., McPhail, M.J.W., 2015. Magnetic Resonance Imaging: Principles and Techniques: Lessons for Clinicians. J Clin Exp Hepatol. 10.1016/j.jceh.2015.08.001

11. Hansson, O., Lehmann, S., Otto, M., Zetterberg, H., Lewczuk, P., 2019. Advantages and disadvantages of the use of the CSF Amyloid β (Aβ) 42/40 ratio in the diagnosis of Alzheimer’s Disease. Alzheimers Res Ther. 10.1186/s13195-019-0485-0

12. Hojjati, S.H., Ebrahimzadeh, A., Babajani-Feremi, A., 2019. Identification of the early stage of alzheimer’s disease using structural mri and resting-state fmri. Front Neurol 10. 10.3389/fneur.2019.00904

13. Imtiaz, B., Tolppanen, A.M., Kivipelto, M., Soininen, H., 2014. Future directions in Alzheimer’s disease from risk factors to prevention. Biochem Pharmacol. 10.1016/j.bcp.2014.01.003

14. Jack, C.R., Bernstein, M.A., Fox, N.C., Thompson, P., Alexander, G., Harvey, D., Borowski, B., Britson, P.J., Whitwell, J.L., Ward, C., Dale, A.M., Felmlee, J.P., Gunter, J.L., Hill, D.L.G., Killiany, R., Schuff, N., Fox-Bosetti, S., Lin, C., Studholme, C., DeCarli, C.S., Krueger, G., Ward, H.A., Metzger, G.J., Scott, K.T., Mallozzi, R., Blezek, D., Levy, J., Debbins, J.P., Fleisher, A.S., Albert, M., Green, R., Bartzokis, G., Glover, G., Mugler, J., Weiner, M.W., 2008. The Alzheimer’s Disease Neuroimaging Initiative (ADNI): MRI methods. Journal of Magnetic Resonance Imaging. 10.1002/jmri.21049

15. Karas, G.B., Scheltens, P., Rombouts, S.A.R.B., Visser, P.J., Van Schijndel, R.A., Fox, N.C., Barkhof, F., 2004. Global and local gray matter loss in mild cognitive impairment and Alzheimer’s disease. Neuroimage 23, 708–716. 10.1016/j.neuroimage.2004.07.006

16. Knopman, D.S., Amieva, H., Petersen, R.C., Chételat, G., Holtzman, D.M., Hyman, B.T., Nixon, R.A., Jones, D.T., 2021. Alzheimer disease. Nat Rev Dis Primers 7. 10.1038/s41572-021-00269-y

17. LaMontagne, P.J., Benzinger, T.L.S., Morris, J.C., Keefe, S., Hornbeck, R., Xiong, C., Grant, E., Hassenstab, J., Moulder, K., Vlassenko, A., Raichle, M.E., Cruchaga, C., Marcus, D., 2019. OASIS-3: Longitudinal Neuroimaging, Clinical, and Cognitive Dataset for Normal Aging and Alzheimer Disease. medRxiv. 10.1101/2019.12.13.19014902

18. Li, W., Wen, W., Chen, X., Ni, B., Lin, X., Fan, W., 2020. Functional Evolving Patterns of Cortical Networks in Progression of Alzheimer’s Disease: A Graph-Based Resting-State fMRI Study. Neural Plast 2020. 10.1155/2020/7839536

19. Littlejohns, T.J., Holliday, J., Gibson, L.M., Garratt, S., Oesingmann, N., Alfaro-Almagro, F., Bell, J.D., Boultwood, C., Collins, R., Conroy, M.C., Crabtree, N., Doherty, N., Frangi, A.F., Harvey, N.C., Leeson, P., Miller, K.L., Neubauer, S., Petersen, S.E., Sellors, J., Sheard, S., Smith, S.M., Sudlow, C.L.M., Matthews, P.M., Allen, N.E., 2020. The UK Biobank imaging enhancement of 100,000 participants: rationale, data collection, management and future directions. Nat Commun. 10.1038/s41467-020-15948-9

20. Miller, K.L., Alfaro-Almagro, F., Bangerter, N.K., Thomas, D.L., Yacoub, E., Xu, J., Bartsch, A.J., Jbabdi, S., Sotiropoulos, S.N., Andersson, J.L.R., Griffanti, L., Douaud, G., Okell, T.W., Weale, P., Dragonu, I., Garratt, S., Hudson, S., Collins, R., Jenkinson, M., Matthews, P.M., Smith, S.M., 2016. Multimodal population brain imaging in the UK Biobank prospective epidemiological study. Nat Neurosci 19, 1523–1536. 10.1038/nn.4393

21. Mirza-Davies, A., Foley, S., Caseras, X., Baker, E., Holmans, P., Escott-Price, V., Jones, D.K., Harrison, J.R., Messaritaki, E., 2022. The impact of genetic risk for Alzheimer’s disease on the structural brain networks of young adults. Front Neurosci 16. 10.3389/fnins.2022.987677

22. Mueller, S.G., Weiner, M.W., Thal, L.J., Petersen, R.C., Jack, C., Jagust, W., Trojanowski, J.Q., Toga, A.W., Beckett, L., 2008. The Alzheimer’s Disease Neuroimaging Initiative.

23. Sendi, M.S.E., Zendehrouh, E., Ellis, C.A., Fu, Z., Chen, J., Miller, R.L., Mormino, E.C., Salat, D.H., Calhoun, V.D., 2023. The link between static and dynamic brain functional network connectivity and genetic risk of Alzheimer’s disease. Neuroimage Clin 37. 10.1016/j.nicl.2023.103363

24. Sendi, M.S.E., Zendehrouh, E., Miller, R.L., Fu, Z., Du, Y., Liu, J., Mormino, E.C., Salat, D.H., Calhoun, V.D., 2021. Alzheimer’s Disease Projection From Normal to Mild Dementia Reflected in Functional Network Connectivity: A Longitudinal Study. Front Neural Circuits 14. 10.3389/fncir.2020.593263

25. Sindi, S., Mangialasche, F., Kivipelto, M., 2015. Advances in the prevention of Alzheimer’s disease. F1000Prime Rep. 10.12703/P7-50

26. Song, Y.H., Yi, J.Y., Noh, Y., Jang, H., Seo, S.W., Na, D.L., Seong, J.K., 2022. On the reliability of deep learning-based classification for Alzheimer’s disease: Multi-cohorts, multi-vendors, multi-protocols, and head-to-head validation. Front Neurosci 16. 10.3389/fnins.2022.851871

27. Stocker, H., Möllers, T., Perna, L., Brenner, H., 2018. The genetic risk of Alzheimer’s disease beyond APOE ε4: systematic review of Alzheimer’s genetic risk scores. Transl Psychiatry. 10.1038/s41398-018-0221-8

28. Tondelli, M., Wilcock, G.K., Nichelli, P., de Jager, C.A., Jenkinson, M., Zamboni, G., 2012. Structural MRI changes detectable up to ten years before clinical Alzheimer’s disease. Neurobiol Aging 33, 825.e25–825.e36. 10.1016/j.neurobiolaging.2011.05.018

29. Weber, C.J., Carrillo, M.C., Jagust, W., Jack, C.R., Shaw, L.M., Trojanowski, J.Q., Saykin, A.J., Beckett, L.A., Sur, C., Rao, N.P., Mendez, P.C., Black, S.E., Li, K., Iwatsubo, T., Chang, C.C., Sosa, A.L., Rowe, C.C., Perrin, R.J., Morris, J.C., Healan, A.M.B., Hall, S.E., Weiner, M.W., 2021. The Worldwide Alzheimer’s Disease Neuroimaging Initiative: ADNI-3 updates and global perspectives. Alzheimer’s and Dementia: Translational Research and Clinical Interventions 7. 10.1002/trc2.12226

30. Yiannopoulou, K.G., Papageorgiou, S.G., 2013. Current and future treatments for Alzheimer’s disease. Ther Adv Neurol Disord. 10.1177/1756285612461679

31. Yu, J.-T., Tan, L., 2015. Lifestyle changes might prevent Alzheimer’s disease. Ann Transl Med 3, 222. 10.3978/j.issn.2305-5839.2015.09.02

